# Phenotypic Age and Neutrophil-to-Lymphocyte Ratio mediate the association between the triglyceride glucose-body mass index and the cardiovascular mortality risk in hypertensive patients

**DOI:** 10.1101/2025.07.20.25331891

**Authors:** Yu Pan, Xinxin Liu, Siming Pan, Shuang Yin, Xu Zhao, kai Zhang, Yinong Jiang, Yan Liu

## Abstract

**Background:** The triglyceride glucose-body mass index (TyG-BMI) has emerged as a novel marker for insulin resistance and metabolic syndrome. However, its relationship with prognosis in hypertensive patients remains uncertain. This study investigates the association between TyG-BMI and all-cause and cardiovascular mortality in hypertensive patients. Additionally, it explores the mediating roles of the neutrophil-to-lymphocyte ratio (NLR) and Phenotypic Age (PhenoAge) in these associations.

**Methods:** Data from 5164 hypertensive adults participating in the National Health and Nutritional Examination Surveys (NHANES) from 1999 to 2018 were analyzed. Mortality details were obtained from the National Death Index (NDI). Restricted cubic spline (RCS) was used to assess potential nonlinear associations, and weighted Cox proportional hazards models assessed the independent association of TyG-BMI with mortality risk. Time-dependent ROC curve analysis evaluated the predictive ability of TyG-BMI for survival. Structural equation modeling (SEM) explored indirect effects of NLR and PhenoAge on mortality associations.

**Results:** Over a median follow-up of 8.33 years, there were 1005 deaths, including 398 from cardiovascular causes. Participants were stratified by TyG-BMI classes using K-means clustering. Higher TyG-BMI levels were associated with increased risks of all-cause (HR 1.28, 95% CI 1.02–1.60, P = 0.03) and cardiovascular mortality (HR 1.62, 95% CI 1.06–2.46, P = 0.025). Time-dependent ROC analysis showed increasing TyG-BMI predictive accuracy for both all-cause and cardiovascular mortality over 10-, 15-, and 20-years, respectively. RCS analysis revealed a U-shaped relationship between TyG-BMI and mortality risk, with an inflection point at TyG-BMI = 239; deviations below or above this threshold significantly impacted mortality risk. SEM identified NLR and PhenoAge as mediators in high-risk hypertensive individuals (TyG-BMI >239).

**Conclusion:** TyG-BMI predicts all-cause and cardiovascular mortality in hypertensive patients, mediated by NLR and PhenoAge. Incorporating TyG-BMI, NLR, and PhenoAge into clinical practice may help improve risk stratification and guide targeted interventions for high-risk individuals.

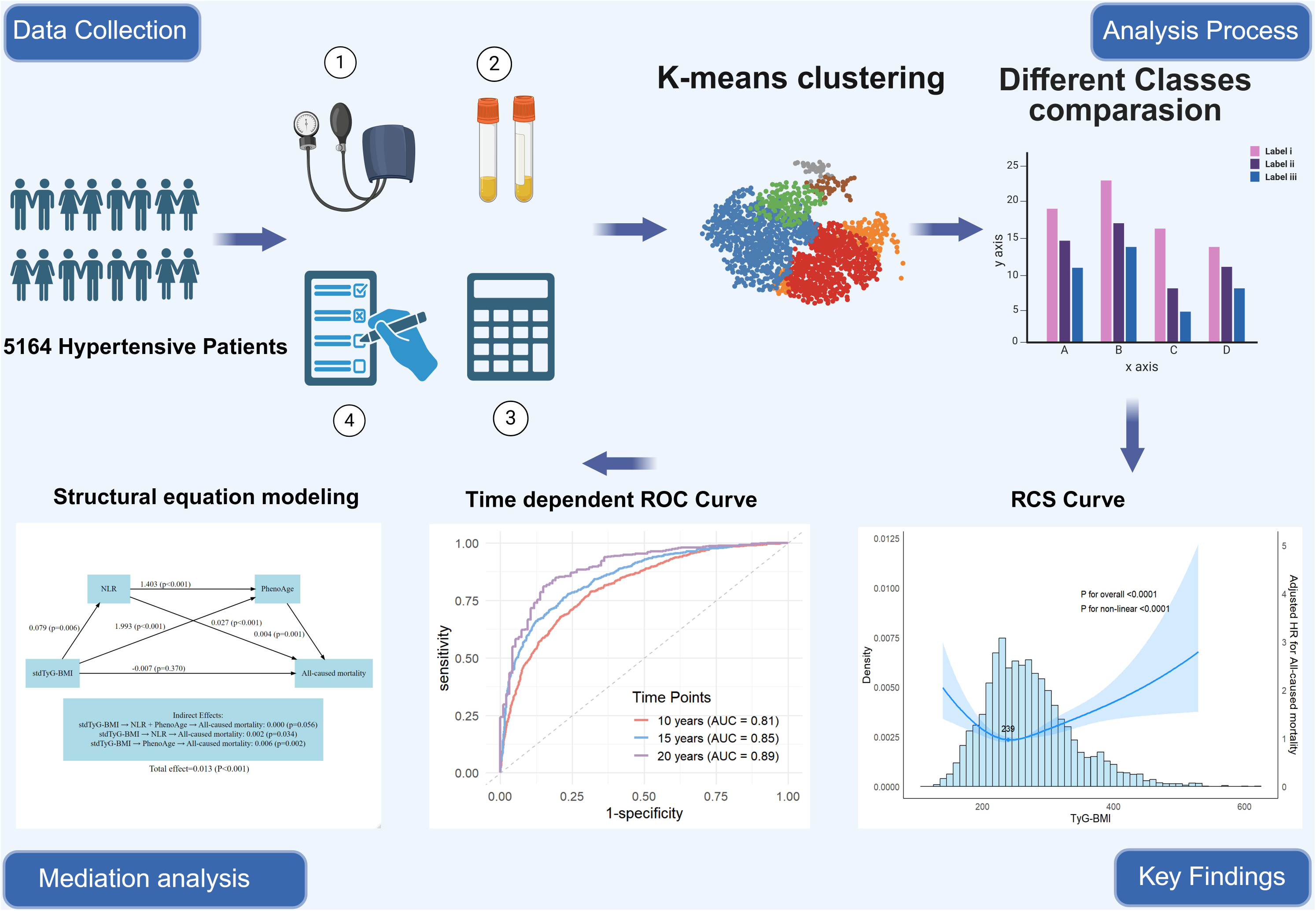

## Introduction

Hypertension represents a significant global public health challenge ^1–3^, affecting an estimated 1.3 billion adults worldwide, with prevalence expected to rise in the forthcoming years^4^. This condition markedly increases the risk of cardiovascular diseases (CVDs), including coronary heart disease, stroke, and heart failure, and is also strongly linked to all-cause mortality^5–7^. The increasing prevalence of hypertension and its poor prognosis are driven by factors such as an aging population and unhealthy lifestyle factors, including high salt and sugar diets and physical inactivity^8,9^.

Recently, the triglyceride-glucose (TyG) index in conjunction with body mass index (BMI) has emerged as a novel marker, referred to as the TyG-BMI, which closely correlates with insulin resistance (IR) and metabolic syndrome^10,11^. These metabolic disturbances are critical factors associated with hypertension and the elevated risk of cardiovascular events^12^. Despite evidence suggesting that TyG-BMI predicts adverse health outcomes ^13,14^, its relationship with mortality in hypertensive individuals remains underexplored.

Moreover, systemic inflammation and biological aging are emerging areas of interest in hypertension research. The neutrophil-to-lymphocyte ratio (NLR) is an established marker of inflammation associated with negative outcomes in hypertensive patients ^15,16^. Concurrently, Phenotypic Age (PhenoAge), which assesses biological age through clinical biomarkers, offers potentially superior predictive power over chronological age^17^. While PhenoAge has been linked to health outcomes in those with IR ^18^, its mediating role alongside NLR in TyG-BMI-related mortality has yet to be thoroughly investigated.

This study aims to bridge the knowledge gap by leveraging data from the National Health and Nutrition Examination Survey (NHANES) from 1999 to 2018. We will investigate the relationship between TyG-BMI and both all-cause and cardiovascular mortality, focusing on hypertensive individuals. Additionally, we will explore the potential mediating roles of NLR and PhenoAge using structural equation modeling (SEM). By examining these interactions, we strive to deepen our understanding of the complex interplay among metabolic dysregulation, inflammation, biological aging, and mortality risk. Our findings aim to offer novel insights and strategies for enhancing long-term survival outcomes in patients with hypertension.

## Methods

### Data source

The information utilized for this research was obtained from the NHANES database from 1999 to 2018, which is managed by the National Center for Health Statistics (NCHS) under the Centers for Disease Control and Prevention (CDC). NHANES represents a nationwide survey designed to collect data regarding the health and nutritional status of civilian residents in the United States. Informed consent was obtained from all participants prior to their involvement in the survey, and the NHANES dataset ensures that identifiable patient information is not included.

### Study population

Blood pressure measurements were conducted by qualified healthcare professionals using a mercury sphygmomanometer at mobile examination centers (MEC). Participants were seated during the measurements, which were primarily taken from the right arm, unless specific circumstances dictated otherwise. After a five-minute resting period, blood pressure readings were obtained consecutively three times, and the average of these readings was used for subsequent analysis. Hypertension was defined as a systolic blood pressure of 140 mmHg or higher, a diastolic blood pressure of 90 mmHg or higher, a self-reported diagnosis of hypertension by a physician, or the use of antihypertensive medication^19^. Participants who lacked complete survival data, blood cell counts, TyG-BMI index, other covariables, and fasting sample weights were excluded from the study. Additionally, individuals with a confirmed diagnosis of cancer or malignant tumors, as well as those with a history of dialysis, were also excluded. Ultimately, the analysis included a total of 5164 participants (Fig. 1).

**Fig. 1.**
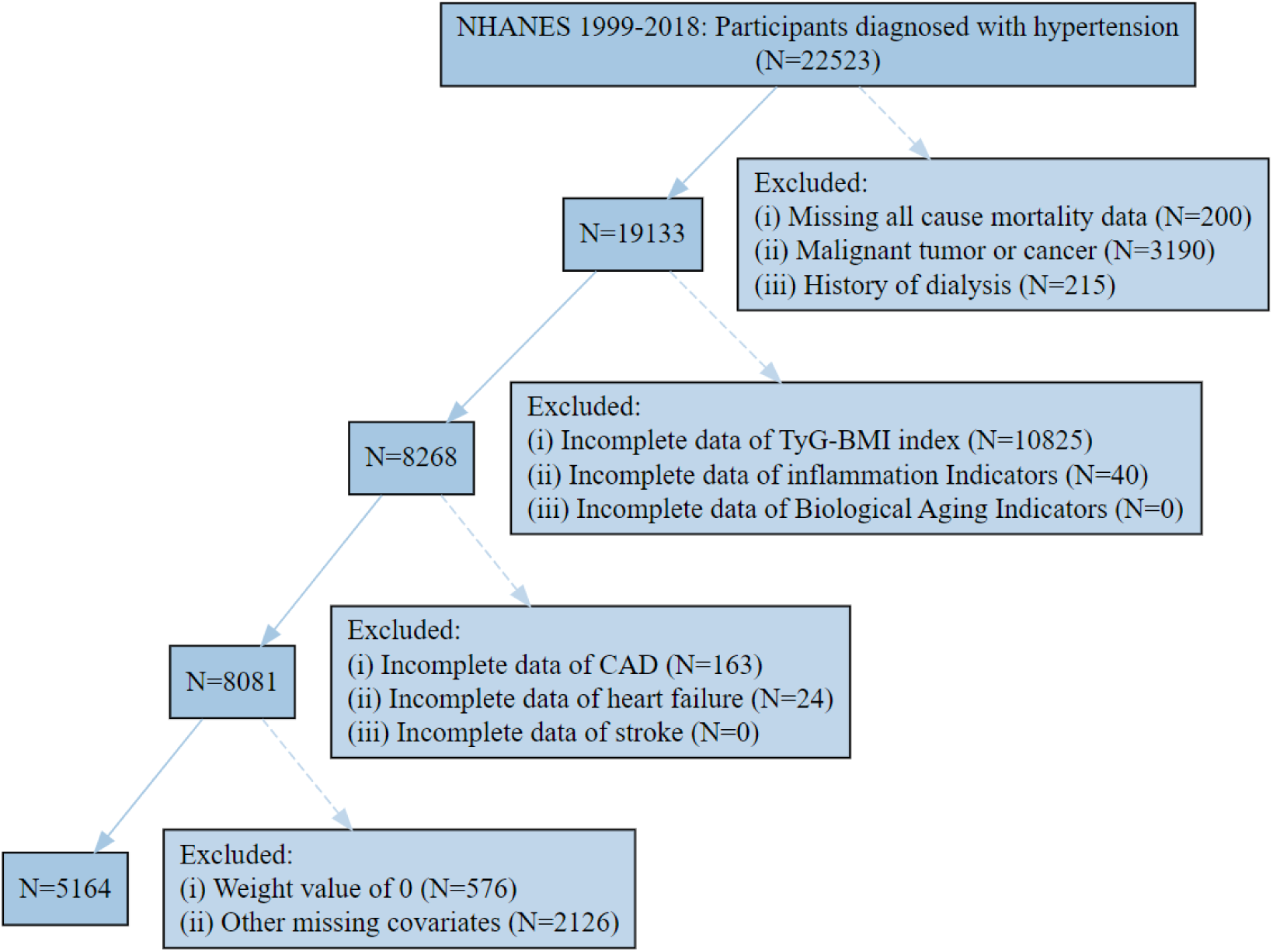
Flowchart of the study population. TyG-BMI: Triglyceride glucose-body mass index; CAD: coronary arterial disease

### Assessment of triglyceride glucose with body mass index

The calculation formula for TyG index is ln [TG (mg/dl) × FPG (mg/dl)/2], and the TyG-BMI index is calculated as TyG×BMI. ^10^.

### Ascertainment of mortality and follow-up

Mortality status was determined by linking NHANES data with records from the National Death Index (NDI), accessible at https://www.cdc.gov/nchs/data-linkage/mortality-public.htm. Participants were classified as either deceased or living based on the information obtained from the NDI. Participants who were not matched with any death records were considered to be alive through the follow-up period. The follow-up duration was calculated from the date of the NHANES examination until either the date of death or December 31, 2019 (updated in 2022), depending on which event occurred first. The underlying causes of death were classified according to the International Classification of Diseases, Tenth Revision (ICD-10). The main outcomes of this study included mortality from all causes and specifically from cardiovascular diseases. Cardiovascular mortality was categorized by the NCHS as deaths associated with heart diseases, identified by ICD-10 codes (I00-I09, I11, I13, I20-I51, I60-I69) ^20^.

### Assessment of inflammation Indicators

The formulas for calculating the NLR, Platelet-to-Lymphocyte Ratio (PLR), Lymphocyte-to-Monocyte Ratio (LMR),Systemic Inflammation Index (SII), and Systemic Inflammation Response Index (SIRI) are as follows: NLR = Neutrophil count / Lymphocyte count, PLR = Platelet count / Lymphocyte count, LMR = Lymphocyte count / Monocyte count, SII = Platelet count × Neutrophil count / Lymphocyte count, and SIRI = Neutrophil count ×Monocyte count / Lymphocyte count^21^.

### Evaluation of Biological Aging Indicators

Indicators of biological aging were evaluated through the Levine method for PhenoAge and the Klemera-Doubal technique for BioAge, both of which utilize clinical laboratory blood chemistry analyses. The BioAge R package was utilized to compute both algorithms^22^. PhenoAge is established through a multivariate analysis approach that combines chronological age with eight specific biomarkers: Albumin, Alkaline phosphatase, Creatinine, Glycated hemoglobin, White blood cell count, Lymphocyte percentage, Mean cell volume, and Red cell distribution width. These biomarkers, together with age, yield a ‘mortality score,’ which is later transformed into a biological age value^23^. In contrast, BioAge is derived using the Klemera-Doubal method, a regression-oriented approach that determines biological age by reducing the discrepancy between the predicted and chronological age within a reference group. This model encompasses nine biomarkers: Forced expiratory volume in 1 second, Systolic blood pressure, Albumin, Alkaline phosphatase, Blood urea nitrogen, Creatinine, C-reactive protein, Glycated hemoglobin, and Total cholesterol^24,25^. Within the BioAge R package, an output will not be generated if more than two of the biomarkers are absent, signifying that while the model can accommodate some missing data, the majority of biomarkers are crucial for precise calculations^26^.

### Ascertainment of covariates

Data pertaining to various demographic and health-related variables were collected, including age, gender, ethnicity, educational level, the ratio of family income to poverty (PIR), BMI, diabetes mellitus, smoking habits, alcohol consumption status, history of cardiovascular disease (CVD), HbA1c, high-density lipoprotein cholesterol (HDL), low-density lipoprotein cholesterol (LDL), total cholesterol (TC), TG, serum creatinine (Scr), serum uric acid (UA), and estimated glomerular filtration rate (eGFR). Race was categorized into Mexican American, Non-Hispanic Black, Non-Hispanic White, and other groups. Education levels were classified as ‘Below high school’ and ‘High School or above’^27^. Smoking status was divided into three categories: never smokers, former smokers, and current smokers^28^. BMI was calculated as weight in kilograms divided by the square of height in meters. According to the diagnostic criteria for diabetes established by the American Diabetes Association, diabetes was defined by the use of insulin or oral hypoglycemic medication, FBG≥126 mg/dL, or an HbA1c level≥6.5%^29^. CVD is defined as a composite of Congestive Heart Failure (CHF), Coronary Artery Disease (CAD), and stroke^30^.The CVD section includes inquiries such as, ‘Has a doctor or other health professional ever informed you that you/he/she had congestive heart failure, coronary heart disease, angina, heart attack, stroke, etc.?’ These questions, labeled as MCQ160B-F in the household questionnaires administered during home interviews, were utilized to identify participants with a history of CVD if they responded ‘yes’ to any of these inquiries. The eGFR was calculated as the following formula: eGFR (mL/min/1.73 m²) = 186 × Scr(-1.154) × Age(-0.203) × 0.742 (if female) ^31^.

### Statistical analyses

The NHANES survey utilizes a complex, multistage, probability sampling design to obtain a nationally representative sample of the non-institutionalized U.S. population. To account for this sampling design and ensure valid statistical inferences, it is essential to apply appropriate sampling weights in the analysis. We utilized the weight of the fasting sample from 1999 to 2018 (WTSAF4YR/ WTSAF2YR) as the new weight. The specific calculation method is as follows: for the years 1999-2002, the new weight is calculated as WTSA4YR × (4 / 20); for the years 2003 and later, the new weight is calculated as WTSAF2YR × (2 / 20).

Data are presented as mean ± standard deviation or proportions. We employed either a weighted χ^2^ test for categorical variables or a weighted linear regression model for continuous variables to assess differences among the various TyG-BMI classes.

We employed the K-means clustering algorithm based on Euclidean distance to categorize patients according to their TyG-BMI values. K-means was selected for its computational efficiency and ease of visualization. This method partitions the data into K clusters by minimizing the within-cluster sum of squares. The procedure involves applying the elbow rule to identify the optimal number of clusters, randomly selecting initial cluster centers, assigning each patient to the nearest centroid, and sequentially updating the centroids. This iterative process continues until the within-cluster sum of squares is minimized. Through cluster analysis, we classified TyG-BMI into three categories: the first category (low level) has a TyG-BMI value of 223 ± 33.8; the second category (moderate level) has a value of 281 ± 43.8; and the third category (high level) has a value of 360 ± 55.1, as illustrated in Figure 2.

**Fig. 2.**
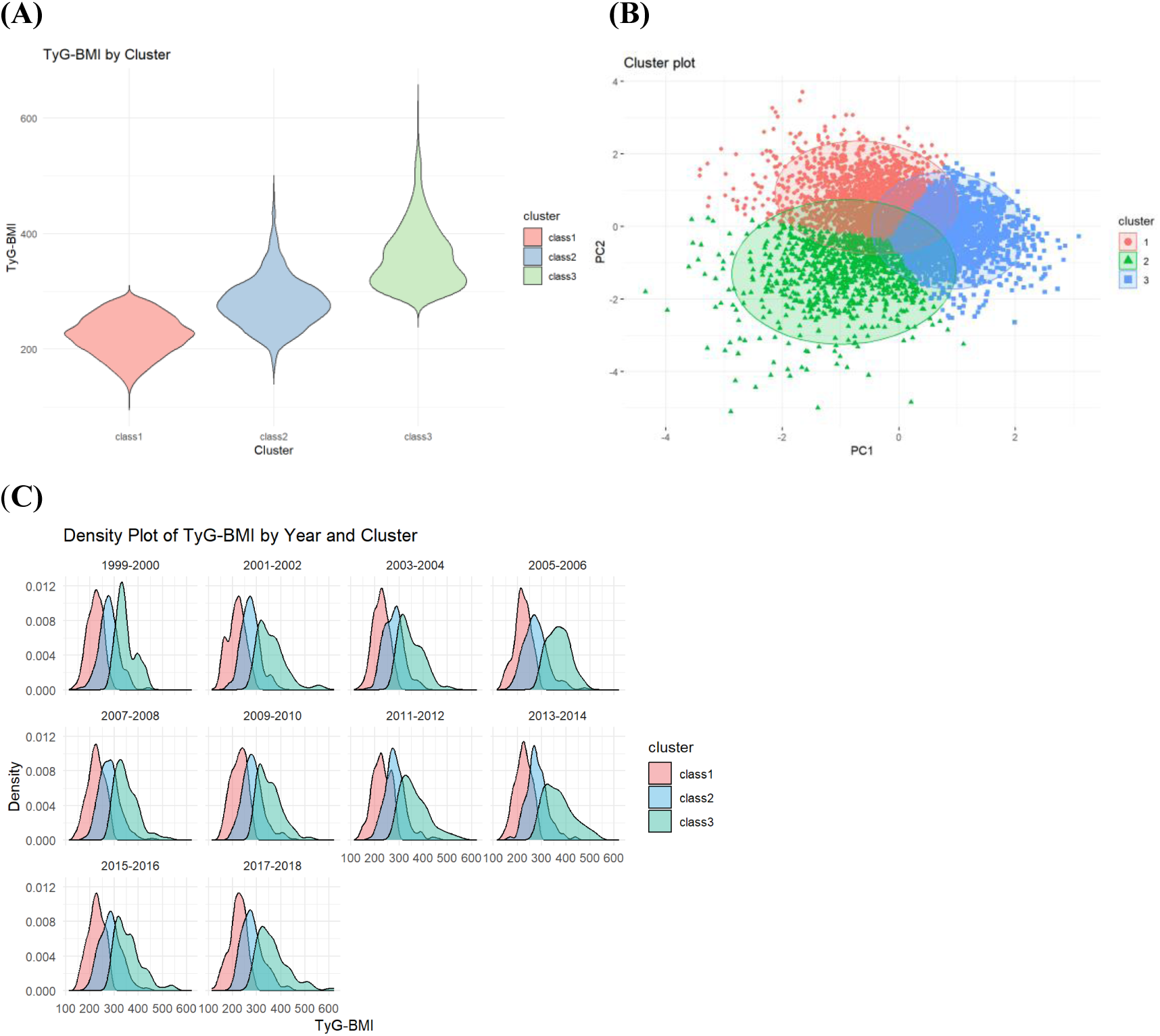
The results of the K-means clustering. (**A)** Three clusters were found using the K-means method with Euclidean distance: the x- and y-axes are principal components of the change in the TyG-BMI; **(B)** Data visualization for the classes of the change in the TyG-BMI: Class 1 (n=2317) exhibited a TyG-BMI value of 223±33.8; Class 2 (n=1869) showed a TyG-BMI value of 281±43.8; Class 3 (n=978) had a TyG-BMI value of 360±55.1; (**C)** The distribution of TyG-BMI classes by years.

We employed restricted cubic splines (RCS) for evaluation. Following the criteria set by the Akaike Information Criterion (AIC), we selected four knots. The shape of the curve derived from the RCS determines the position of the inflection point.

Cox proportional hazards models, weighted by survey data, were utilized to evaluate the independent relationship between TyG-BMI and both all-cause and cardiovascular mortality among patients with hypertension. The ‘timeROC’ package was utilized to assess the accuracy of TyG-BMI in predicting survival outcomes at different time points^15,32^. Stratified and interaction analyses were conducted based on variables such as age (<60 years vs. ≥60 years), sex (male vs. female), race (Mexican American, non-Hispanic Black, non-Hispanic White, and other), and eGFR (< 90 vs. ≥90 mL/min/1.73 m²).

In this study, we employed SEM to investigate the impact of a TyG-BMI value greater than 239 on all-cause mortality and cardiovascular mortality. Analyses were conducted using the lavaan package in R. Our hypothesis posits that TyG-BMI influences all-cause mortality or cardiovascular mortality in hypertensive patients through two mediating variables: NLR and PhenoAge. Simultaneously, we adjusted for multiple potential confounders in the model, including age, gender, race, smoking status, alcohol consumption, marital status, education level, and PIR. Specific methods for SEM can be referenced in the supplementary materials. Statistical analyses were performed using R version 4.4.2 and R studio version 2024.9.1.394. A two-sided p-value of less than 0.05 was considered statistically significant.

## Results

### Baseline characteristics of study participants

This study included a total of 5,164 participants who were categorized into three groups based on TyG-BMI levels using cluster analysis. The baseline characteristics and intergroup differences are presented in Table 1. Participants in Class 3, which represented the group with the highest TyG-BMI levels, had the youngest average age and the highest proportion of female patients, Paradoxically, aging-related indicators, such as phenoAge and bioAge, were the lowest in this group, despite the presence of elevated cardiovascular and inflammatory risks factors. This group also had the lowest proportion of individuals who had never smoked or consumed alcohol. Additionally, class 3 demonstrated the highest eGFR values, indicating better renal function. However, obesity-related indicators, such as BMI and abdominal circumference, exhibited the highest values in Class 3, along with elevated levels of inflammatory markers, such as SII and SIRI. Regarding cardiovascular diseases, the incidence of stroke was the highest.

**Table 1.**
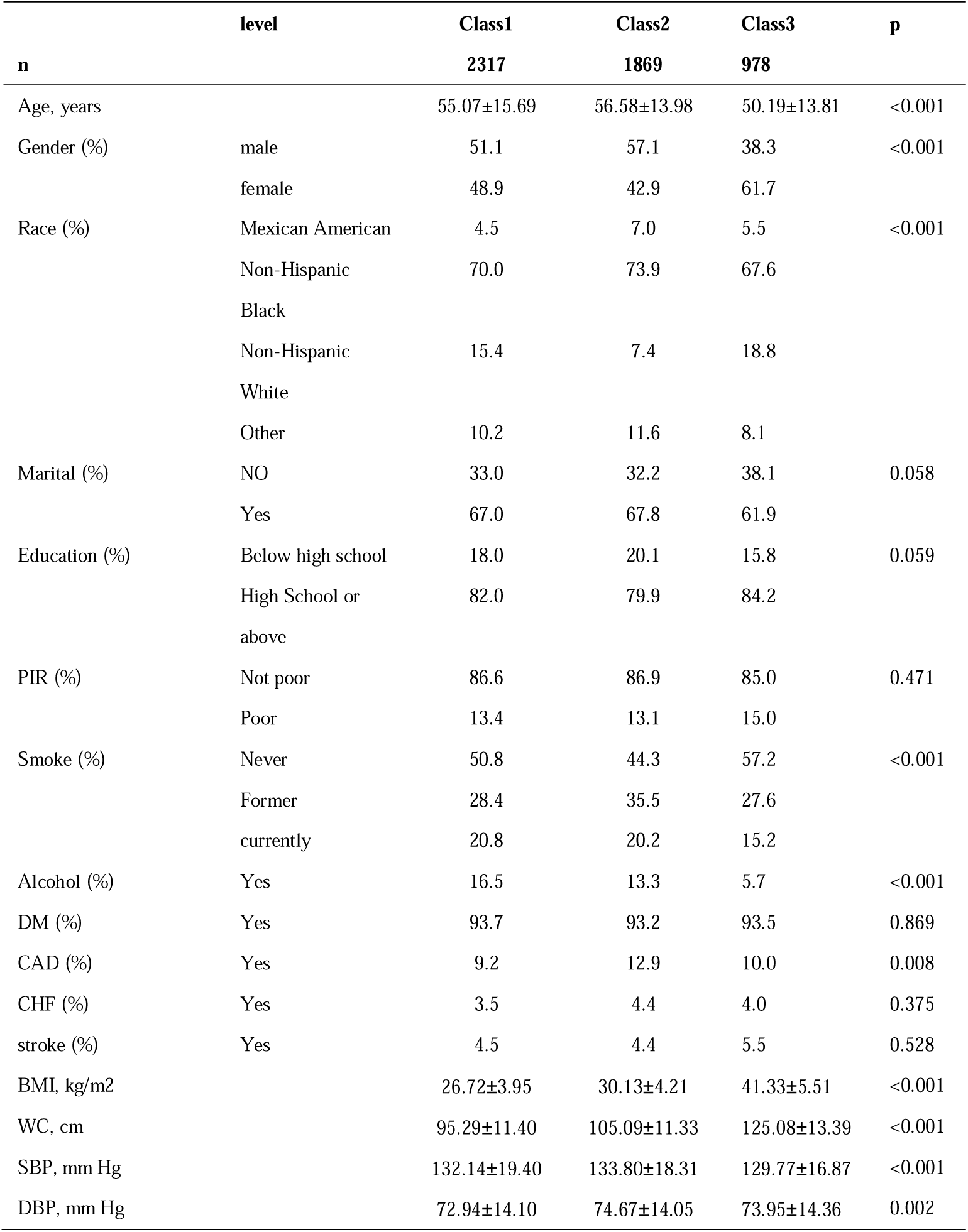
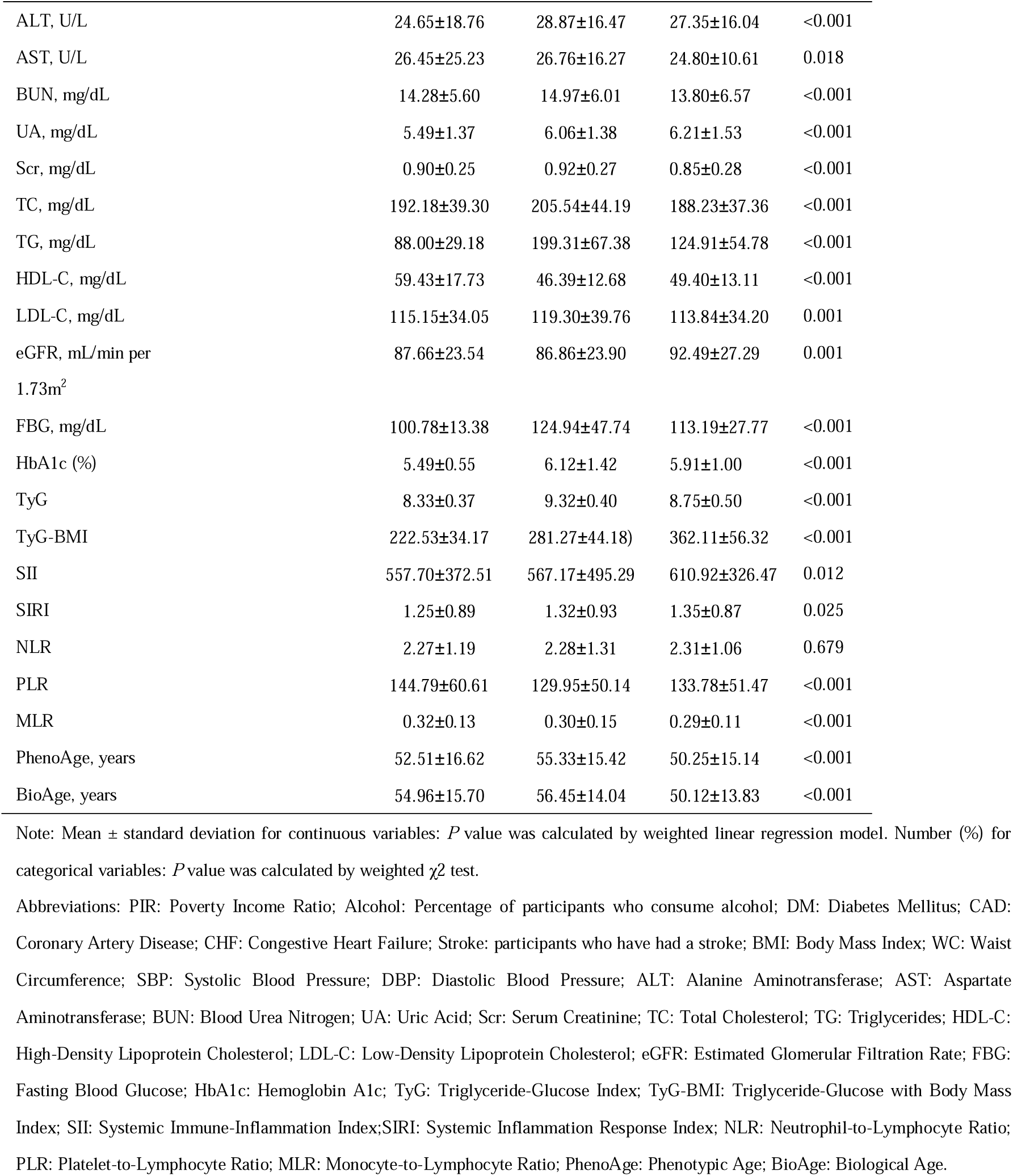
Baseline characteristics of study participants.

### Associations of TyG-BMI with all-cause and cardiovascular mortality in patients with hypertension

The median duration of follow-up was 8.33 years, with an interquartile range of 4.58 to 12.42 years. Over 5,164 person-years of follow-up, there were 1,005 deaths from all causes, of which 398 were linked to CVD. We developed three models to assess the independent impact of TyG-BMI classes on mortality. The hazard ratios (HRs) and 95% confidence intervals (CIs) for the three classifications are detailed in Table 2. In the completely adjusted model (Model 3), we categorized TyG-BMI according to the findings from the cluster analysis. In comparison to individuals in Class 1 (low TyG-BMI), those classified in Class 3 (high TyG-BMI) showed a significantly higher risk of all-cause mortality, with an HR of 1.28 (95% CI, 1.02–1.60). Furthermore, participants in both Class 2 (moderate TyG-BMI) and Class 3 demonstrated a significantly increased risk of cardiovascular mortality, with HRs of 1.36 (95% CI, 1.04–1.78) and 1.62 (95% CI, 1.06–2.46), respectively.

**Table 2.** The relationships between different TyG-BMI classes and mortality risk.

|  | **Model 1** |  | **Model 2** |  | **Model 3** |  |
| --- | --- | --- | --- | --- | --- | --- |
|  | **OR (95% CI)** | **P value** | **OR (95% CI)** | **P value** | **OR (95% CI)** | **P value** |
| All-caused mortality |  |  |  |  |  |  |
| Class1 | Reference |  | Reference |  | Reference |  |
| Class2 | 1.24(1.04,1.48) | 0.017 | 1.16(0.98,1.39) | 0.091 | 1.17(0.99,1.39) | 0.072 |
| Class3 | 0.76(0.61,0.93) | 0.009 | 1.26(1.01,1.58) | 0.041 | 1.28(1.02,1.60) | 0.03 |
| Cardiovascular mortality |  |  |  |  |  |  |
| Class1 | Reference |  | Reference |  | Reference |  |
| Class2 | 1.37(1.08,1.73) | 0.009 | 1.36(1.04,1.77) | 0.023 | 1.36(1.04,1.78) | 0.023 |
| Class3 | 0.89(0.61,1.29) | 0.5 | 1.56(1.02,2.37) | 0.039 | 1.62(1.06,2.46) | 0.025 |

Model 1, unadjusted; Model 2, adjusted for age, sex, race, marital, education level, the ratio of family income to poverty, smoking status, alcohol consumption and Diabetes Mellitus; Model 3, adjusted for age, sex, race, marital, education level, the ratio of family income to poverty, smoking status, alcohol consumption and Diabetes Mellitus, eGFR, SBP and DBP.

Subgroup analyses were conducted to further explore the association between TyG-BMI and both all-cause and cardiovascular mortality risks across various demographic and clinical variables, including age, gender, race, and eGFR. The results indicated that the effect of TyG-BMI on all-cause and cardiovascular mortality was consistent across all subgroups, with no significant interactions effects observed (*P* for interaction > 0.05) (Supplementary Table 1).

### Nonlinear association of TyG-BMI with all-cause and cardiovascular mortality in patients with hypertension

To explore the potential nonlinearity between TyG-BMI and mortality, we utilized a Cox proportional hazards regression model featuring restricted cubic splines. The smooth curve fitting, fully adjusted for age, sex, race, marital, education level, the ratio of family income to poverty, smoking status, alcohol consumption, Diabetes Mellitus, eGFR, SBP, and DBP, indicated a nonlinear U-shaped relationship between TyG-BMI and both all-cause and cardiovascular mortality (Fig. 3). Furthermore, we assessed the link between TyG-BMI and mortality through both a Cox proportional hazards model and a piecewise two-part Cox proportional hazards model. As indicated in Table 3, the inflection point for TyG-BMI was identified at a value of 239. For TyG-BMI values below this threshold, a one standard deviation increase in TyG-BMI was significantly correlated with a lower risk of all-cause mortality (HR = 0.53; 95% CI, 0.38-0.72; *P* < 0.001) and a similarly reduced risk of cardiovascular mortality (HR = 0.28; 95% CI, 0.32-0.80; *P* = 0.004). Conversely, for TyG-BMI values surpassing 239, a one standard deviation increase was significantly associated with a heightened risk of all-cause mortality (HR = 1.22; 95% CI, 1.07-1.38; *P* = 0.003) and cardiovascular mortality (HR = 1.24; 95% CI, 1.04-1.48; *P* = 0.018).

**Fig. 3.**
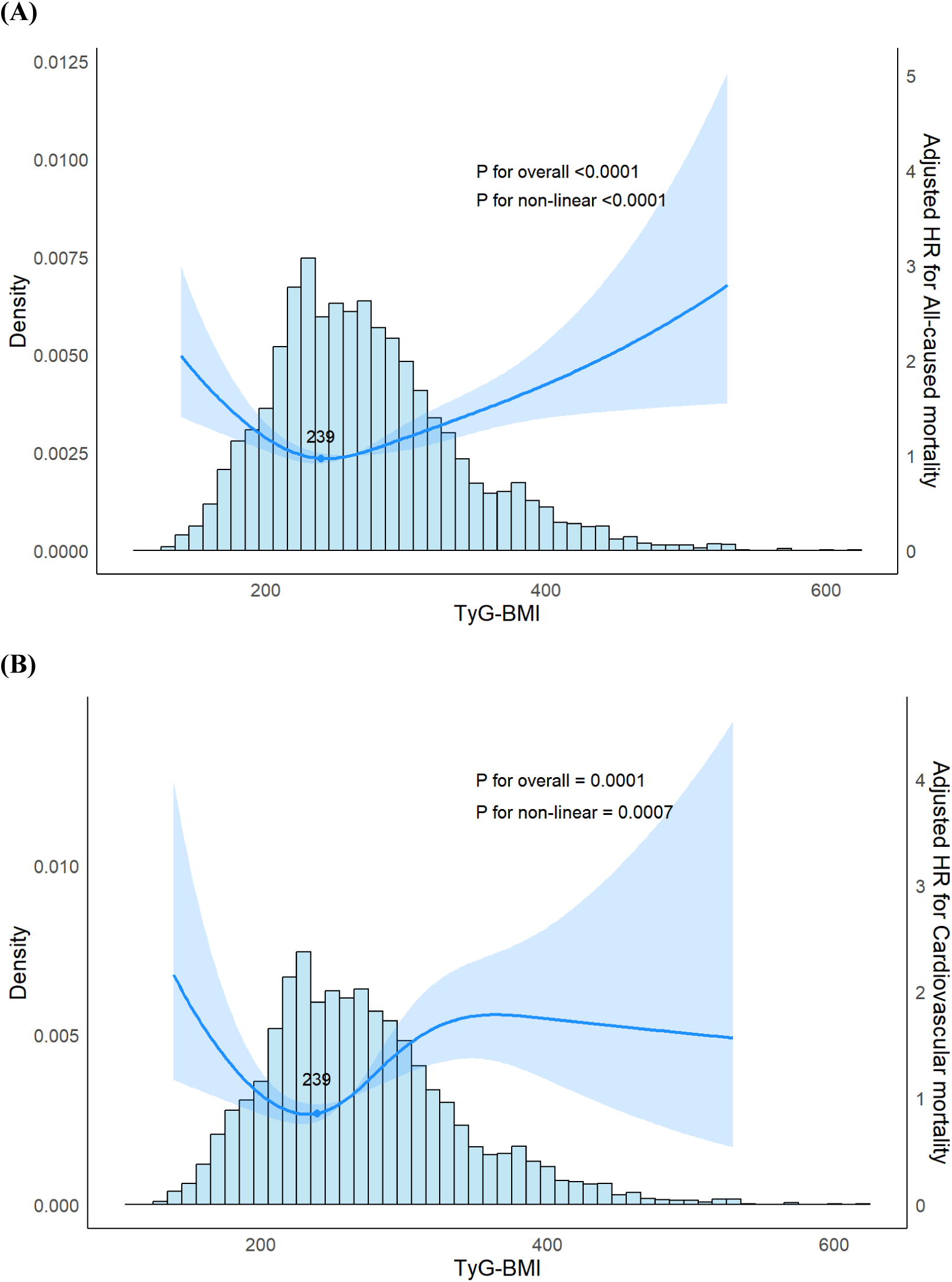
The association of TyG-BMI with mortality among hypertension visualized by restricted cubic spline. **(A)** All-cause mortality; **(B)** Cardiovascular mortality. Hazard ratios were adjusted for age, sex, race, marital, education level, the ratio of family income to poverty, smoking status, alcohol consumption and Diabetes Mellitus, eGFR, SBP and DBP.

**Table 3.**
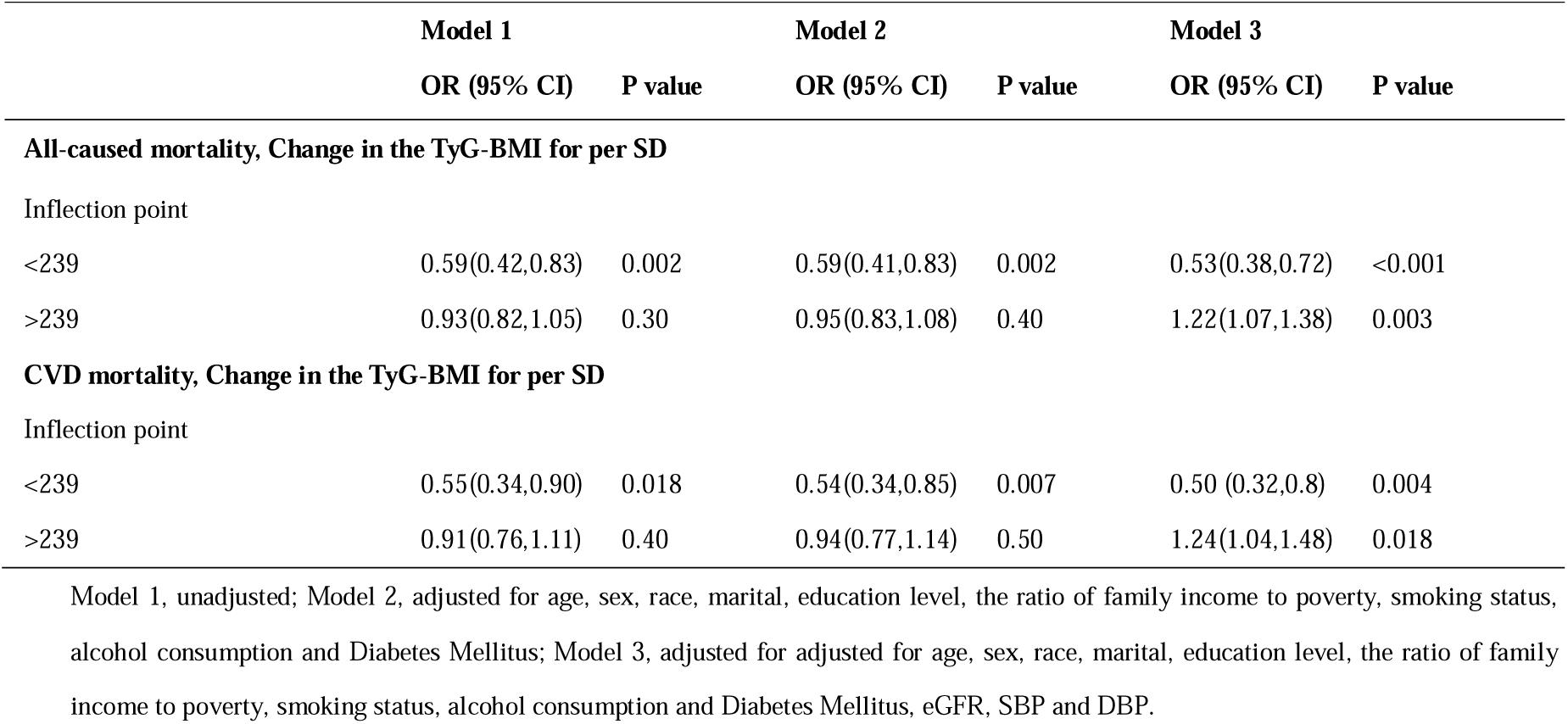
A piecewise two-part Cox proportional hazards model.

### The predictive ability of TyG-BMI for all-cause and cardiovascular mortality in patients with hypertension

Time-dependent ROC curve analysis revealed that AUC for the TyG-BMI was 0.81, 0.85, and 0.89 for 10-year, 15-year, and 20-year all-cause mortalities, respectively (Fig. 4A). Similarly, the AUC for TyG-BMI was 0.83, 0.87, and 0.91 for 10-year, 15-year, and 20-year cardiovascular mortalities, respectively (Fig. 4B). These results demonstrate the strong predictive ability of TyG-BMI for both all-cause and cardiovascular mortality over time.

**Fig. 4.**
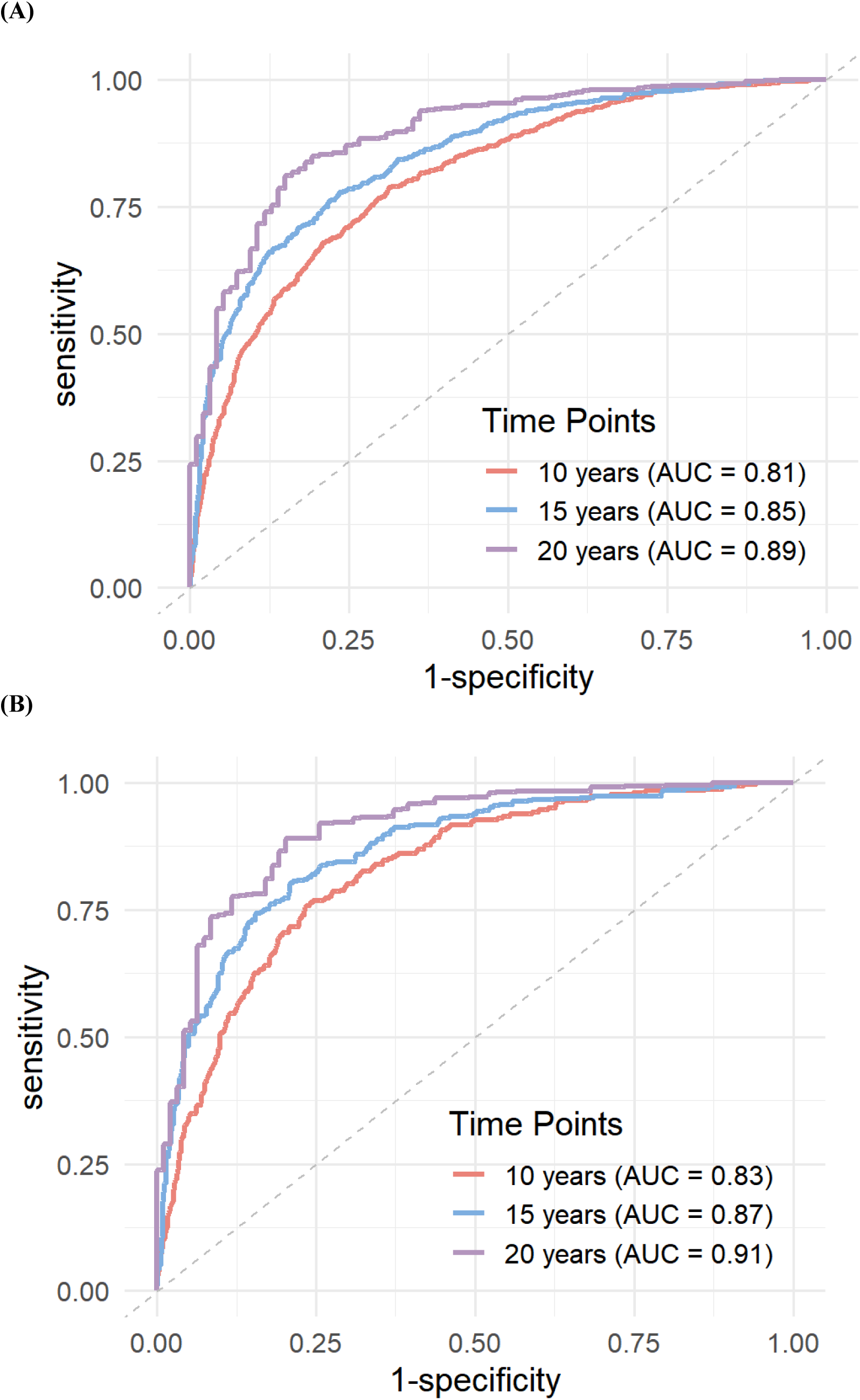
Time-dependent ROC curves of the NLR for predicting all-cause mortality. **(A)** All-cause mortality; **(B)** Cardiovascular mortality. The ROC curves were adjusted for age, sex, race, marital, education level, the ratio of family income to poverty, smoking status, alcohol consumption and Diabetes Mellitus, eGFR, SBP and DBP.

### Mediation analysis

In the SEM analysis focused on individuals with a TyG-BMI greater than 239 (n=3326), we observed significant indirect effects of TyG-BMI on all-cause mortality, mediated through PhenoAge and NLR. Specifically, the indirect effect of TyG-BMI on all-cause mortality through NLR was 0.002 (*P* =0.034), while the indirect effect through PhenoAge was 0.006 (*P* =0.002). The total effect of TyG-BMI on all-cause mortality was estimated to be 0.013 (*P* <0.001). Model fit indices demonstrated good performance, with CFI=0.995, TLI=0.963, and RMSEA=0.061. The indirect effect of TyG-BMI on cardiovascular mortality through NLR alone was not significant (0.001, *P* =0.194), whereas the indirect effect of TyG-BMI on cardiovascular mortality through PhenoAge alone was significant, estimated at 0.004 (*P* =0.003). The total effect of TyG-BMI on cardiovascular mortality was estimated to be 0.008 (*P* =0.001). Model fit indices demonstrated strong performance in this context (CFI=0.997, TLI=0.977, RMSEA=0.047), and the effect values of each mediation path are illustrated in Fig. 5(A, B).

**Fig. 5.**
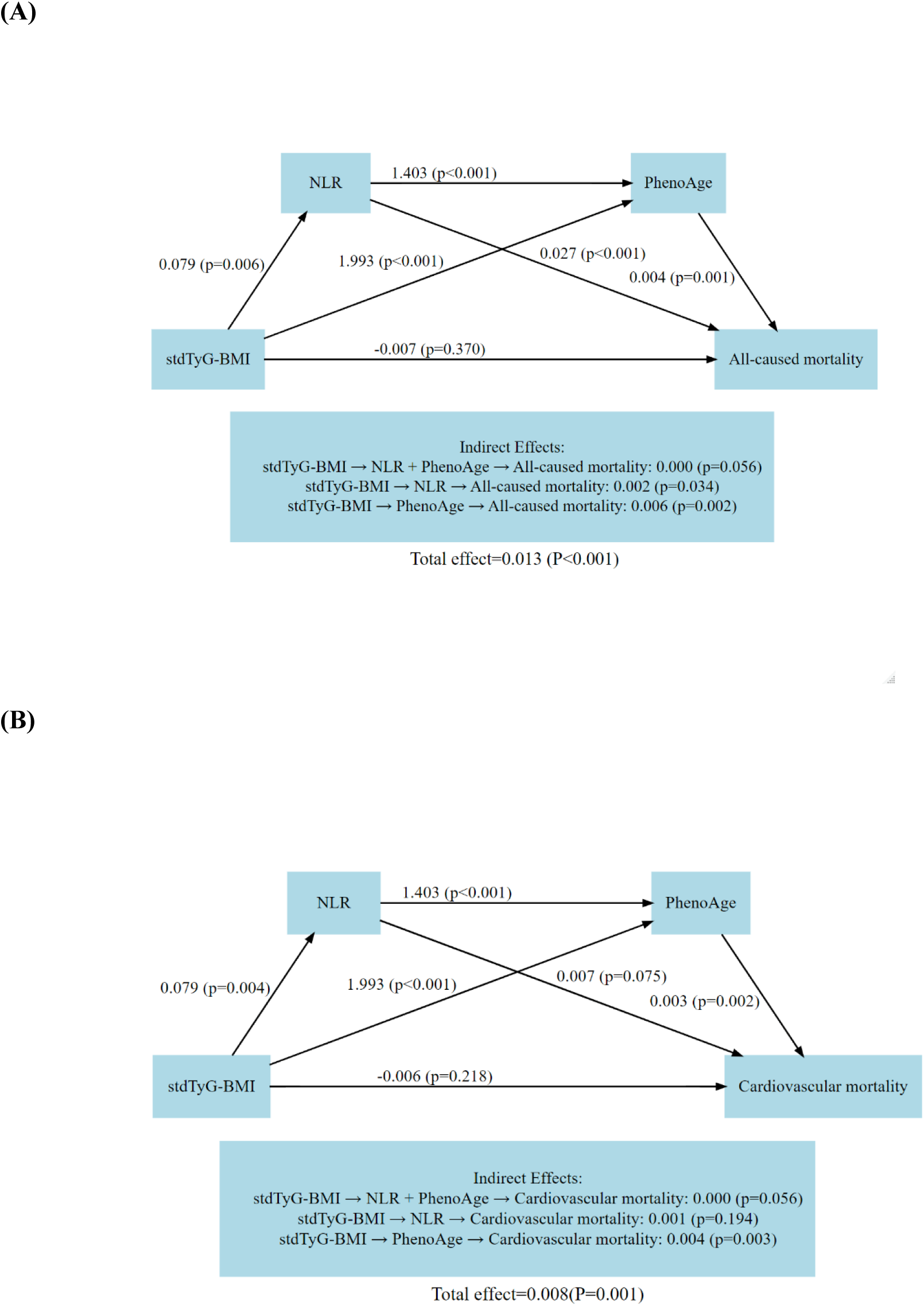
Sequential Chain Mediation Analysis Using Structural Equation Modeling. **(A)** All-cause mortality; **(B)** Cardiovascular mortality. stdTyG-BMI: Standardized Triglyceride-Glucose Body Mass Index. We combined the method of principal component analysis (PCA) to help adjust for confounding factors such as age, gender, race, smoking status, alcohol consumption, marital status, education level, and PIR, while also helping to reduce model complexity and dimensionality.

## Discussion

In this analysis of 5,164 hypertensive patients based on the NHANES database, we identified a significant association between TyG-BMI and both all-cause and cardiovascular mortality, with TyG-BMI demonstrating robust, time-dependent predictive power. Specifically, hypertensive individuals with a TyG-BMI greater than 239 exhibited a notably higher mortality risk. Additionally, our findings revealed that NLR and PhenoAge mediated the relationship between TyG-BMI and mortality risk, further elucidating the mechanisms underlying this association.

Obesity is widely recognized as one of the most prominent risk factors for the development of hypertension^33^.The interplay between obesity and IR exacerbates the pathophysiology of hypertension, exerting a synergistic effect on blood pressure regulation^34^. The TyG index, derived from triglycerides and glucose levels, has emerged as a readily accessible indicator of insulin sensitivity and a recognized risk marker for IR^35,36^. On the other hand, BMI, calculated from height and weight, is commonly utilized to assess the risk of obesity and metabolic diseases^37,38^. Recent studies have indicated that the TyG-BMI index, which incorporates TyG index and BMI, is superior to the TyG index or BMI alone in predicting IR. Comparative analyses of the TyG-BMI index with other related indicators, such as the TyG index combined with waist circumference (TyG-WC) or the waist-to-height ratio (TyG-WHtR), have indicated that the TyG-BMI index outperforms these other markers in predicting IR^10,39^. Moreover, TyG-BMI index has been linked to several adverse health outcomes, including ischemic stroke^14,40^, cardiovascular mortality risk in diabetic patients^41^, and the incidence and prognosis of cardiovascular disease^41,42^. Previous study has also suggested an association between TyG-BMI index and mortality risk in patients with hypertension or coronary heart disease^43^. The current study extends these findings by demonstrating that TyG-BMI index is a significant predictor of both all-cause and cardiovascular mortality in patients with hypertension. Importantly, our study is the first to illustrate that the predictive accuracy of TyG-BMI improves over time. This time-dependent association likely reflects the cumulative effects of metabolic abnormalities and IR, which progressively worsen over the course of hypertension. The results from time-dependent ROC curve underscore the importance of recognizing these cumulative metabolic abnormalities, as they have a profound impact on the predictive model’s performance.

The NLR is a straightforward and reliable indicator of systemic inflammatory status^44^. Elevated NLR levels are often observed with increasing obesity grades, reflecting a heightened inflammatory response, particularly in individuals with chronic low-grade inflammation^45^. Inflammation is widely recognized as a critical factor in the onset and progression of atherosclerosis. Previous studies have indicated that hypertensive patients are particularly prone to low-grade chronic inflammation, which accelerates arteriosclerosis and increases the risk of cardiovascular events^46,47^. This study demonstrated that NLR significantly mediated the relationship between TyG-BMI and both all-cause and cardiovascular mortality. This suggested that metabolic abnormalities not only elevated mortality risk directly, but also amplified it through the promotion of systemic inflammatory responses. Elevated NLR levels in individuals with high TyG-BMI further reinforced the strong link between metabolic syndrome and systemic inflammation. These results align with the findings of Xuexue Zhang et al. (2024), who reported a positive association between NLR and cardiovascular risk in hypertensive patients^15^. Notably, our study advances this understanding by clarifying the mediating role of NLR in the pathway between metabolic indicators, such as TyG-BMI, and mortality risk. The mediating effect of NLR highlights the critical role of inflammation in exacerbating the adverse outcomes associated with metabolic syndrome and hypertension. Targeting systemic inflammation through pharmacological or lifestyle interventions may thus represent an effective approach to reducing cardiovascular and all-cause mortality in hypertensive patients with high TyG-BMI.

PhenoAge, a measure of a “biological age”, is calculated using a series of biomarkers that provide a more accurate reflection of an individual’s health status and mortality risk compared to chronological age. An increased PhenoAge typically indicates accelerated biological aging and a heightened risk of diseases^25^. Previous research has demonstrated that hypertension contributes to accelerated biological aging, potentially influencing PhenoAge through mechanisms such as oxidative stress, inflammation, and metabolic disorders^48^. While the association between biological aging and adverse cardiovascular outcomes is well-documented, the underlying mechanisms remain unclear^49^. Our findings indicate that PhenoAge serves as a mediator in the relationship between TyG-BMI and mortality, suggesting that metabolic disorders increase the mortality risk not only via traditional cardiovascular risk factors but also by hastening the biological aging process. This highlights a novel pathway by which metabolic disorders, including insulin resistance and obesity, may contribute to increased mortality, independent of established cardiovascular risks. Moreover, the NLR not only independently increase mortality risk, but also promotes cellular senescence, exacerbating the biological aging process^50^. During the aging process, a phenomenon known as the ‘Senescence-Associated Secretory Phenotype (SASP)’ occurs, wherein senescent cells release a substantial array of inflammatory factors, including chemokines, proteases, and other substances^51^. The release of these SASP factors aggravates both local and systemic inflammatory responses, further promoting the senescence of neighboring cells and perpetuating a vicious cycle^52^. In the context of hypertension and elevated TyG-BMI, this chronic inflammatory environment, marked by elevated NLR, accelerates systemic aging and impairs local tissue. This bidirectional feedback loop between inflammation and biological aging significantly elevates mortality risk in patients with hypertension, particularly in those with a higher TyG-BMI index.

The U-shaped relationship observed in the RCS curve indicates that both high and low TyG-BMI values are associated with increased mortality risk. For individuals with a low TyG-BMI, several potential factors may contribute to this elevated risk: 1. Malnutrition: A low BMI can be indicative of insufficient nutritional intake, which may weaken immune function and increase vulnerability to infections and other diseases^53^; 2. Metabolic disorders: An excessively low TyG index may suggest that the body has been in a prolonged low energy state, adversely affecting insulin sensitivity and metabolic balance; 3. Underlying diseases: Several chronic conditions can lead to reductions in body weight and lipid levels, indicating that low TyG-BMI may be a marker for these underlying health issues; 4. Insufficient physiological reserves: Lower body weight often correlates with reduced physiological reserves, limiting the body’s ability to cope with acute health events, such as trauma or surgery.

The primary strengths of this study lie in its status as the first systematic investigation of the association between the TyG-BMI index and mortality in patients with hypertension, along with a comprehensive analytical approach utilizing advanced statistical methods such as RCS, weighted Cox proportional hazards models, and SEM. SEM is a robust multivariate analysis tool capable of simultaneously addressing multiple path relationships and accounting for various confounding factors, thereby providing more comprehensive insights into the complex relationships between TyG-BMI, inflammation, biological aging and mortality. Our study contributes to the existing literature by uncovering the intricate mediating effects within these associations, suggesting novel potential biomarkers, such as NLR and phenoAge, for early intervention and risk stratification in patients with hypertension. Ultimately, understanding these mechanisms may guide the development of more targeted treatment strategies aimed at reducing mortality risk throughout the course of hypertension. Additionally, time-dependent ROC curve analysis demonstrated the robust predictive ability of TyG-BMI for all-cause and cardiovascular mortality at different time points, underscoring its potential utility in clinical risk stratification.

### Limitations

While we possess follow-up data characteristic of cohort studies, the NHANES dataset lacks detailed information regarding the grade of hypertension, which may constrain the findings. Additionally, the study population is drawn solely from the United States, which may restrict the generalizability of the results to other populations. Another limitation is that the SEM-based sequential chained mediation analysis focused only on individuals with a TyG-BMI exceeding 239, which does not adequately explain the adverse impact of excessively low TyG-BMI values on outcomes. In this context, although significant efforts were made to adjust for confounding factors, further external validation is necessary to confirm these findings and enhance their applicability.

### Conclusion

This study highlights the mediating roles of NLR and PhenoAge in the relationship between TyG-BMI and mortality, shedding light on the complex mechanisms by which metabolic syndrome contributes to increased mortality risk in patients with hypertension. These findings not only reinforce the intricate interplay among metabolism, inflammation, and biological aging, but also offers new insights and potential biomarkers for future clinical intervention strategies.

## Data Availability

The data used for these analyses are all publicly available at online (https://www.cdc.gov/nchs/nhanes/index.htm).

## Author Contributions

YL and YJ designed the study. YP conducted the primary research and performed data analysis and wrote the manuscript. XL and SP assisted with data analysis, drafted the results part and helped the manuscript revisions. SY assisted with data collection and interpretation. XZ assisted with data analysis. KZ contributed to the literature review and manuscript revisions. All authors had full access to all the data in the study. All authors reviewed and approved the final manuscript and had final responsibility for the decision to submit for publication.

## Acknowledgements

We highly appreciate the work by participants in the NHANES project.

## Ethics approval and consent to participate

The study was approved by the National Center for Health Statistics NCHS Research Ethics Review Board. All participants signed the informed consent before participating in this survey.

