## Supplement Table 1 for "Phenotypic Age and Neutrophil-to-Lymphocyte Ratio mediate the association between the triglyceride glucose-body mass index and the cardiovascular mortality risk in hypertensive patients"

Supplementary Table1 Subgroup analysis of the associations between TyG-BMI and mortality

| Characteristics | All-cause mortality | | | P interaction | Cardiovascular mortality | | | P interaction |
|---|---|---|---|---|---|---|---|---|
| | **Class1** | **Class2** | **Class3** | | **Class1** | **Class2** | **Class3** | |
| | | **HR (95%CI), *P*** | **HR (95%CI), *P*** | | | **HR (95%CI), *P*** | **HR (95%CI), *P*** | |
| **Age** | | | | 0.55 | | | | 0.90 |
| <60 | Ref | 1.31 (0.82 - 2.10), 0254 | 1.60 (1.05 - 2.46), 0.031 | | Ref | 1.10 (0.73 - 2.29), 0.258 | 1.37 (0.77 - 2.93), 0.422 | |
| >60 | Ref | 0.86 (0.52 - 1.42), 0.551 | 0.69 (0.39 - 1.24), 0.221 | | Ref | 0.95 (0.45 - 2.01), 0.899 | 0.77 (0.29 - 2.10), 0.615 | |
| **Sex** | | | | 0.12 | | | | 0.28 |
| Male | Ref | 1.22 (0.95 - 1.56), 0.117 | 1.58 (1.09 - 2.31), 0.017 | | Ref | 0.63 (0.44 - 0.92), 0.017 | 0.63 (0.28 - 1.41), 0.260 | |
| Female | Ref | 0.92 (0.63 - 1.33), 0.647 | 0.67 (0.41 - 1.11), 0.119 | | Ref | 1.38 (0.80 - 2.34), 0.247 | 1.49 (0.82 - 2.93), 0.195 | |
| **Race** | | | | 0.68 | | | | 0.67 |
| Mexican American | Ref | 1.45 (0.89 - 2.35), 0.135 | 1.22 (0.71 - 2.10), 0.479 | | Ref | 0.88 (0.46 - 1.70), 0.708 | 1.20 (0.50 - 2.85), 0.685 | |
| Non-Hispanic Black | Ref | 1.01 (0.64 - 1.59), 0.961 | 1.20 (0.51 - 2.82), 0.682 | | Ref | 1.32 (0.78 - 2.24), 0.299 | 1.87 (0.59 - 5.91), 0.286 | |
| Non-Hispanic White | Ref | 1.60 (0.95 - 2.69), 0.078 | 1.09 (0.44 - 2.73), 0.848 | | Ref | 2.18 (1.10 - 4.31), 0.025 | 1.00 (0.29 - 3.50), 0.999 | |
| Other | Ref | 1.41 (0.70 - 2.84), 0.332 | 1.14 (0.34 - 3.85), 0.833 | | Ref | 1.22 (0.32 - 4.66), 0.773 | 1.79 (0.33 - 9.60), 0.497 | |
| **eGFR** | | | | 0.15 | | | | 0.78 |
| <90 | Ref | 0.90 (0.60 - 1.35), 0.613 | 1.51 (0.86 - 2.62), 0.148 | | Ref | 0.91 (0.49 - 1.71), 0.775 | 2.34 (0.89 - 6.14), 0.083 | |
| >90 | Ref | 1.25 (0.87 - 1.80), 0.226 | 0.98 (0.64 - 1.52), 0.932 | | Ref | 1.48 (0.86 - 2.53), 0.157 | 0.90 (0.42 - 1.93), 0.784 | |

HR were adjusted for age, sex, race, marital, education level, the ratio of family income to poverty, smoking status, alcohol consumption and Diabetes Mellitus, eGFR, SBP and DBP.

**Supplementary Materials**

In this study, we employed SEM to investigate the impact of a TyG-BMI value greater than 239 on all-cause mortality and cardiovascular mortality. Analyses were conducted using the lavaan package in R. Our hypothesis posits that TyG-BMI influences all-cause mortality or cardiovascular mortality in hypertensive patients through two mediating variables: NLR and PhenoAge. Simultaneously, we adjusted for multiple potential confounders in the model, including age, gender, race, smoking status, alcohol consumption, marital status, education level, and PIR.To simplify the model's complexity and reduce data dimensionality, we first standardized the data, including the TyG-BMI variable. Subsequently, we performed principal component analysis (PCA) on the categorical variables. The number of extracted principal components was determined based on the cumulative variance ratio, leading to the selection of the first two principal components (PC1 and PC2) for further analysis. These principal components account for most of the variance in the dataset, effectively reducing the model's dimensionality. In our statistical inference, we applied NHANES sample weights to enhance the generalizability and robustness of the results. When constructing the SEM model, we focused on three indirect effect paths: the mediating effects on all-cause mortality or cardiovascular mortality mediated by NLR and PhenoAge, as well as the total effect, which illuminated the complex relationships among the variables. To enhance the reliability of our inference results and facilitate their interpretation, we utilized 1,000 bootstrap samples to estimate the model parameters, thereby providing robust effect estimates and p-values.
